# Detection and characterization of single SARS-CoV-2 viral particles by flow virometry

**DOI:** 10.64898/2026.04.28.26351941

**Authors:** Martin Jungbauer-Groznica, Pierre-Henri Commere, Cyril Planchais, Andrea Cottignies-Calamarte, Alejandro De Cruz, Aura Fantin Rengifo, Florence Guivel-Benhassine, Isabelle Staropoli, Sandrine Schmutz, Sophie Novault, David Veyer, Hélène Péré, Hugo Mouquet, Olivier Schwartz, Timothée Bruel

## Abstract

Virus infected cells release viral particles, which have variable protein content and are functionally diverse. Deciphering this heterogeneity remains a challenge. Here, we adapt flow virometry to detect and phenotype severe acute respiratory syndrome coronavirus 2 (SARS-CoV-2) particles. In supernatants of infected cells, we observe particles measuring 70–100 nm. The appearance of these particles is associated to the increase in viral RNA and infectivity. Sample inactivation using temperature or detergent leads to the disappearance of these particles. Using antibodies and dyes for lipid membranes and nucleic acids, we detect the spike protein, the lipid envelope and the RNA genome. We further confirm the presence of viral particles by electron microscopy. Analyzing different viral preparations demonstrate that spike detection in particles outcompetes particle concentration to predict infectivity. Antibodies against different spike epitopes enable probing of spike conformation changes in the presence of soluble ACE2. Lastly, we detect SARS-CoV-2 particles in PCR-confirmed patient nasal swabs without prior purification steps. In summary, we developed an efficient framework to detect and characterize single SARS-CoV-2 particles.

## Introduction

Any given viral population, being in a patient sample or in a culture dish, is not a uniform entity but a population of genetically, and phenotypically diverse particles [1]. Multiple mechanisms underly this heterogeneity. Transcriptional errors during replication create variation in genetic content, stochastic events during particle assembly generate differences in protein content (either of viral or host origin) or particle size, and, in the case of enveloped viruses, the localization of viral budding may change lipid composition. Some viral species also undergo extracellular maturation processes or produce particles of difference shapes, which further add to the phenotypic and functional diversity. This pleomorphic nature of viruses has a direct impact on infectiousness and immune evasion [2]. However, investigations have thus far been largely restricted to particles with distinct morphologies or carrying defective genomes.

One of the main barriers for studying viral particles is their small size. Conventional techniques such as ELISA or electron microscopy are either restricted to bulk analyses or low throughput. Next-generation sequencing has unlocked the study of defective genomes with unprecedented detail and mass spectrometry has unraveled the protein composition of particles, but neither achieves single-viral-particle characterization. Additionally, many techniques don’t use authentic virus (e.g. pseudovirus) or involve selective purification steps (enrichment/depletion of certain populations), which likely alter the composition of the viral preparation. Finally, existing methods are difficult to apply to patient samples. The technological advancement in nanoscale flow cytometry made it possible to resolve particles smaller than 300–500 nm and thus lifted many limitations for the direct study of viral particles, also known as flow virometry (FVM) [3]. By utilizing light and fluorescence scattering, FVM is a sensitive, multiparametric, high-throughput method for the detection, quantification, and characterization of individual viral particles [4]. Yet, despite promising preliminary validations, the extent to which FVM consistently fulfils these expectations across different viral species and contexts (i.e. culture supernatants and patient samples) requires further systematic evaluation. Moreover, it remains notably underutilized for the detection and phenotyping of SARS-CoV-2 or other coronaviruses.

Severe acute respiratory syndrome coronavirus 2 (SARS-CoV-2), the causative agent of coronavirus disease 2019 (COVID-19), and human or zoonotic coronaviruses in general are enveloped, negative-strand RNA viruses with a size of approximately 70–100 nm and [5]. Since the beginning of the pandemic, SARS-CoV-2 has continued to evolve giving rise to numerous variants that notably contain numerous mutations in the spike glycoprotein (S) [6]. Being crucial for receptor interaction and fusion, the alterations in the different spike domains have had lasting consequences on tissue tropism, replication kinetics, and especially antibody immune evasion. Cryo-EM revealed that the number of S is variable within viral particles, with functional consequences that are ill-defined [7]. Studies have also elaborated on the incorporation of host proteins within SARS-CoV-2 virions. For instance, proteomics analyses found the presence of G3BP1/2 and CD59, two major stress granule nucleators and a potent inhibitor of the complement membrane attack complex, respectively [8]. Still, how the incorporation and the variation of viral and host proteins within viral particles shape viral fitness remains poorly defined. Filling this gap would require combining single viral particles analysis with functional assay, which is currently technically challenging.

Here, we adapted FVM for the detection and phenotyping of SARS-CoV-2 particles. In infected cell supernatants, we identify 70-100 nm particles that increase with viral RNA and infectivity and that are absent in uninfected cells. We design stainings for the detection of spike protein, genomic RNA and lipid envelope. By analyzing different SARS-CoV-2preparations, we highlight spike incorporation as a correlate of infectivity. We show that changes in spike conformation can be monitored by antibodies targeting different spike domains. Lastly, we detect spike-positive SARS-CoV-2 particles in PCR-confirmed patient nasal swabs without purification steps.

## Materials and Methods

### Ethics statement and patient samples

Our study was retrospective and informed consent from all participants was obtained after collection of the samples. Our observational work was carried out in accordance with the Declaration of Helsinki with no sampling addition to usual procedures. Swab specimens were obtained for standard diagnostic following medical prescriptions in our hospital. The project was approved by the ethics committee “Comite d’ethique de la recherche AP-HP Centre“ affiliated to the AP-HP (Assistance publique des Hopitaux de Paris). Nasal swabs were collected in the two cavities and preserved in 3 ml of transport M4RT buffer (Remel Microtask, Thermo Fisher).

### Cell lines and viral isolates

U2OS-angiotensin-converting enzyme 2 (U2OS-ACE2) cells stably expressing the GFP (green fluorescent protein) split system (GFP1-10 and GFP11; called S-Fuse cells), IGROV-1 and VeroE6 cells have been described previously [9, 10]. Cells were grown in dulbecco’s modified eagle medium containing GlutaMAX™ (DMEM-GlutaMAX) and supplemented with 10% fetal calf serum (FCS) and 1% penicillin-streptomycin (PS), later refer to as “complete medium”, at 37°C and 5% CO_2_. Blasticidin (10 mg/mL) and puromycin (1 mg/mL) were used to select for ACE2 and GFP transgene expression, respectively. Absence of mycoplasma contamination was confirmed with the Mycoalert Mycoplasma Detection Kit (Lonza). SARS-CoV-2 isolates D614G and XBB.1.5 have been described previously and were prepared on VeroE6 and IGROV-1 cells respectively and titrated with the S-Fuse assay [11–13].

### D614G kinetics of progeny virus in supernatant

One day before the experiment, VeroE6 cells (5×10^5^ per well) were plated in 6-well plates. The next day, cells were washed with phosphate-buffered saline (PBS) and incubated with D614G in conditioned medium (DMEM-GlutaMAX and 1% PS, without FCS) at MOI 0.1 and 0.001 respectively for 1 hour at 37°C. After incubation, cells were washed with PBS and medium was added for further incubation at 37°C. For time point 0 hours post infection (hpi), the supernatant was collected immediately for background baseline value while other time points were collected as indicated (1, 6, 24 and 48 hours). Collected supernatants were centrifugated at 21,310 x g for 15 minutes at 4°C and stored at -80°C for subsequent RT-qPCR, infectivity and FVM analyses. Cells were detached, washed in PBS, fixed in 4% paraformaldehyde and stored at 4°C for flow cytometry analyses.

### Viral RNA quantification with RT-qPCR

Samples containing SARS-CoV-2 virus were heat-inactivated for 20 minutes at 80°C. Ten μM of SARS-CoV-2 E-gene Forward (5′-ACAGGTACCTTAATAGTTAATAGCGT-3′) and Reverse (5′-ATATT GCAGCAGTACGCACACA-3′) primers was used with Luna Universal One-step RT-qPCR Kit (New England Biolabs) and added to 1 μL supernatant (5 μL total) in a 384-well plate. A standard curve was produced through a 1:10 serial dilution of EURM-019 ssRNA SARS-CoV-2 fragments for reference (European Commission). RT-qPCR was carried out using iTaq universal SYBR green supermix (BioRad). All RT-qPCR experiments were performed with a QuantStudio 6 Flex Real-Time PCR machine.

### Virus titration with S-Fuse assay

U2OS-angiotensin-converting enzyme 2 (U2OS-ACE2) cells harboring GFP1-10 or GFP11 (called S-Fuse cells) become GFP+ when they are productively infected with SARS-CoV-2 [9, 11]. Cells were plated (ratio 1:1; 2×10^4^ cells per well total) in black, glass-bottom µClear 96-well plates (Greiner Bio-One). Viral samples were added to the S-fuse cells at 1:5 serial dilutions for 18 hours at 37°C 5% CO2, fixed with 2% paraformaldehyde, washed with PBS and stained with Hoechst (dilution 1:1,000, Invitrogen H3570). Images were acquired on an Opera Phenix high content confocal microscope (PerkinElmer). The GFP area and the number of nuclei were quantified using the Harmony software (PerkinElmer). The viral titer (infectious units /mL; IU) was calculated from the last positive dilution with 1 infectious unit (IU) being 3 times the background (GFP area in non-infected controls). We previously reported correlations between infectious unit obtained with the S-Fuse assay and TCID50 measured in VeroE6 cells [14].

### Flow cytometry of SARS-CoV-2-infected cells

SARS-CoV-2-infected cells were intracellularly stained with anti-SARS-CoV-2 nucleoprotein (N) antibody (clone NCP-1, [10])[10]) conjugated to DyLight-488 (2 μg/mL) in PBS with 0.05% saponin, 1% BSA and 0.05% sodium azide for 30 minutes at room temperature. After the staining, the cells were washed with PBS and acquired on an Attune Nxt instrument (ThermoFisher). Data was analyzed with FlowJo V10.10.0 (BD Life Science).

### Flow virometry

#### Instrument calibration

For the analyses of SARS-CoV-2 particles, two machines were used: the Nanoflow Analyzer (NanoFCM) and CytoFLEX nano (Beckman).

For the Flow NanoAnalyzer, the calibration of the optics (filter and laser) was performed with polystyrene beads (250 nm diameter; NanoFCM) diluted (1:100) in deionized water filtered using a Whatman Anotop syringe filter (25 mm diameter, 0.02 µm porosity). They also served as concentration standards to determine the particles per ml. Excitation was provided by a 488 laser (9/50 kW) and data collected through a 488 ± 5 nm bandpass filter for side scatter and 525 ± 40 nm bandpass filters for fluorescence. The size determination of particles was based on the side scatter intensities of silica nanosphere sizing standard set (S16M-Exo; 68 nm, 91 nm, 113 nm, 155 nm; NanoFCM) excited at 488 nm and measured through the 488 ± 5 nm filter. Standard curves were constructed from side scatter intensities and experimental particle diameter was determined in the already included software. All quality control and experimental samples were acquired at the sampling pressure of 1kPa for 1 minute with 2,000 to 12,000 events per minute. The thresholding was performed using the automatic threshold capability of the included software, which was set to “small signal”. Data was acquired with the NF Profession software (versions 2.0) and further analyzed with FlowJo V10.10.0 (BD Life Science).

For the CytoFLEX nano, the calibration of the optics (filter and laser) was performed with polystyrene nanoparticles (144 nm diameter; Beckman C85323) and fluorescence polystyrene nanoparticles (500 nm diameter; Beckman C85324). Calibration of the fluorescence sensitivity was performed with multi-fluorescence polystyrene nanoparticles (500 nm diameter; Beckman C92889). Excitation was provided by four lasers: 405 nm (120 mW), 488 nm (50 mW), 561 (35 mW) and 638 nm (100 mW) and data collected through bandpass filters 447±60, 531±46, 595±50 and 670±30 for fluorescence and 405±10, 488±8, 561±6 and 638±6 for scatter. Sample line and sample tube were acquired for 30 seconds at 1 µl/min flow rate with a first gate (P1; set to less than 150 events on a control tube of PBS) aiming at eliminating the background noise of the sheath fluid and of the PBS (0.02 µm filtered) used to dilute the sample. Actual samples were acquired at 3 µl per minute with an event rate of approximately 5000 events per second. Data was acquired with the CytExpert software (versions 2.7) and further analysed with FlowJo V10.10.0 (BD Life Science). For the size determination of particles, data was further processed using the FCMpass software (https://www.fcmpass.com/#) [15–17]. Slightly different sizes are displayed in the data. This is due to different versions of FCMpass that were used to calibrate the size. In version 5.0.7, we used the parameter “EV Diameter (nm) [High RI]” (refractive index (RI) of 1.406) while in version 5.0.13, we used “Enveloped virus” (refractive index (RI) of 1.456).

#### Staining of viral samples

For staining of SARS-CoV-2 particles, cell culture supernatants were incubated with Tixagevimab conjugated to DyLight-488 (1 µg/mL) or Syto24 (20 µM; Thermo Fisher S7559) or MemGlow-488 (200 nM; Cytoskeleton #MG01) for 30 minutes at room temperature (RT). For the determination of particle swarming, virus was first serially diluted before being stained with Tixagevimab conjugated to DyLight-488.

For the probing of spike confirmation, virus was incubated with anti-spike monoclonal antibodies (mAbs; 1 µg/mL) and Syto24 (20 µM) and 90 µg/mL of soluble ACE2 (produced in-house) for 30 minutes at room temperature before adding anti-human IgG Alexa Fluor 647 (AF647; 2 µg/mL).

For the detection of SARS-CoV-2 particles in nasal swabs, virus containing buffer was incubated with SA55 conjugated to DyLight-488 (1 µg/mL) for 15 minutes at room temperature. After the staining, the sample was fixed with paraformaldehyde (4% final) and diluted in Dulbecco’s phosphate-buffered saline (PBS; Gibco) to match with manufacturers’ instruction for optimal flow rate (*i.e* number of events per second). Antibody-fluorophore conjugations were performed with the DyLight™ 488 NHS Ester kit (Thermo Fisher 46403) in order to avoid the usage of secondary antibodies and thus reducing fluorescent background [18].

### Transmission electron microscopy (TEM)

Standard 200 mesh formvar coated and carbon stabilized grids were discharged for 15 seconds with 2 mA at 0.1 mBar using a Quorum Q150 ES unit. 3 μl of sample was pipetted onto the activated grid and incubated for 3 minutes at room temperature before the grid was washed three times in Mili-Q water for 30 seconds respectively and finally stained with 2% uranyl acetate for 1 minute at room temperature. After staining, grids were dried using Whatman paper. Negative stain images were acquired using a Tecnai T12 BioTWIN 120 kV with a Rio16 detector.

### Monoclonal antibodies (mAbs)

Sotrovimab was provided by CHU Orléans. All other mAbs targeting different SARS-CoV-2 spike epitopes were produced as following. Codon-optimized synthetic DNA fragments coding for the immunoglobulin variable domains of Tixagevimab, SA55, CoV44-62, 76E1 and mGO53 were synthetized (GeneArt, Thermo Fisher Scientific) and cloned into human IgG1 expression or into vectors harboring the LALA and E430G mutations respectively as previously described [19]. Recombinant IgG1 antibodies were produced by transient co-transfection of Freestyle^TM^ 293-F suspension cells (Thermo Fisher Scientific) using the polyethylenimine (PEI) precipitation method and purified from culture supernatants by affinity chromatography using Protein G Sepharose® 4 Fast Flow (GE Healthcare) as previously described [19]. A summary of the mAbs is displayed in **Table 1**.

**Table 1.**
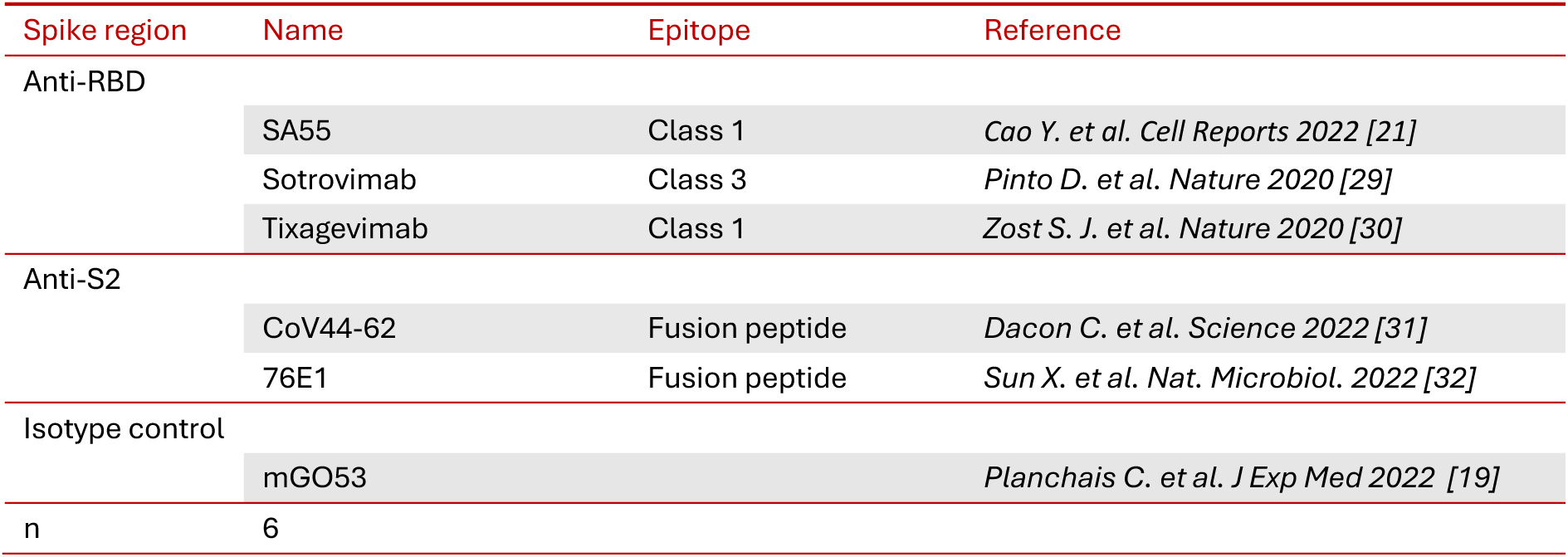
Monoclonal antibodies used for the probing of D614G spike conformations.

| Spike region | Name | Epitope | Reference |
| --- | --- | --- | --- |
| Anti-RBD | SA55 | Class 1 | *Cao Y. et al. Cell Reports 2022 [21]* |
|  | Sotrovimab | Class 3 | *Pinto D. et al. Nature 2020 [29]* |
|  | Tixagevimab | Class 1 | *Zost S. J. et al. Nature 2020 [30]* |
| Anti-S2 | CoV44-62 | Fusion peptide | *Dacon C. et al. Science 2022 [31]* |
|  | 76E1 | Fusion peptide | *Sun X. et al. Nat. Microbiol. 2022 [32]* |
| Isotype control |  |  |  |
|  | mGO53 |  | *Planchais C. et al. J Exp Med 2022 [19]* |
| n | 6 |  |  |

### Statistical analysis

Data processing and calculations were performed with Excel 365 (Microsoft). Figures and statistical analysis were performed using GraphPad Prism Version 10.5.0 (GraphPad software). Statistical significance between different groups was calculated using the tests indicated in each figure legend. Shapiro-Wilk analysis was used to check for normal distribution of data for parametric testing. The correlation analyses used a non-parametric Spearman correlation.

## Results

### Label-free detection and quantification of SARS-CoV-2 particles in crude supernatant

We first asked whether we could detect SARS-CoV-2 particles in supernatant of infected cells using flow virometry (FVM) without any purification or staining steps. We used the Flow NanoAnalyzer (NanoFCM), which has an integrated size estimation tool based on side-scatter intensities. We infected Vero E6 cells at two MOIs (0.1 and 0.001) collected the supernatant at different hours post infection (hpi) and froze them at -80°C. Once all supernatants were collected, we added PFA for fixation. While the supernatant from non-infected cells (NI) showed a modest particle increase between 40–60 nm at both 24 and 48 hpi, the D614G-infected conditions exhibited a stronger elevation of particles between 70–90 nm, but only at 48 hpi (highlighted in red) **(Figure 1A)**. The overall concentration measurement (based on a standard during machine calibration) of all three conditions showed no significant difference in particle count **(Figure 1B)**. However, when gated on the 70–90 nm peak, the infected conditions displayed a significant increase in particle concentration **(Figure 1C)**. The appearance of the 70–90 nm peak was temporally associated with a rise in viral RNA, infectious virions and infected cells, strongly suggesting that these were SARS-CoV-2 progeny virions **(Figure 1D, 1E and 1F)**. Heat-inactivation (80°C for 30 min) or the addition of detergent (1% Triton X-100) led to the disappearance of the 70–90 nm peak **(Figure 1G)**. We also analyzed a reference D614G viral stock (D614G 973) by FVM and observed the same sharp peak localizing at 81.7 ± 13 nm (in line with cryo-EM data of SARS-CoV-2 ; 82 ± 13 (69–95) nm; [5]), while the corresponding non-infected Vero E6 supernatant had a higher particle count in the lower size range **(Figure 1H)**. Thus, SARS-CoV-2 progeny particles can be detected and quantified in supernatants based on their side-scattering features, where they localize around 82 nm, without any purification or staining.

**Figure 1.**
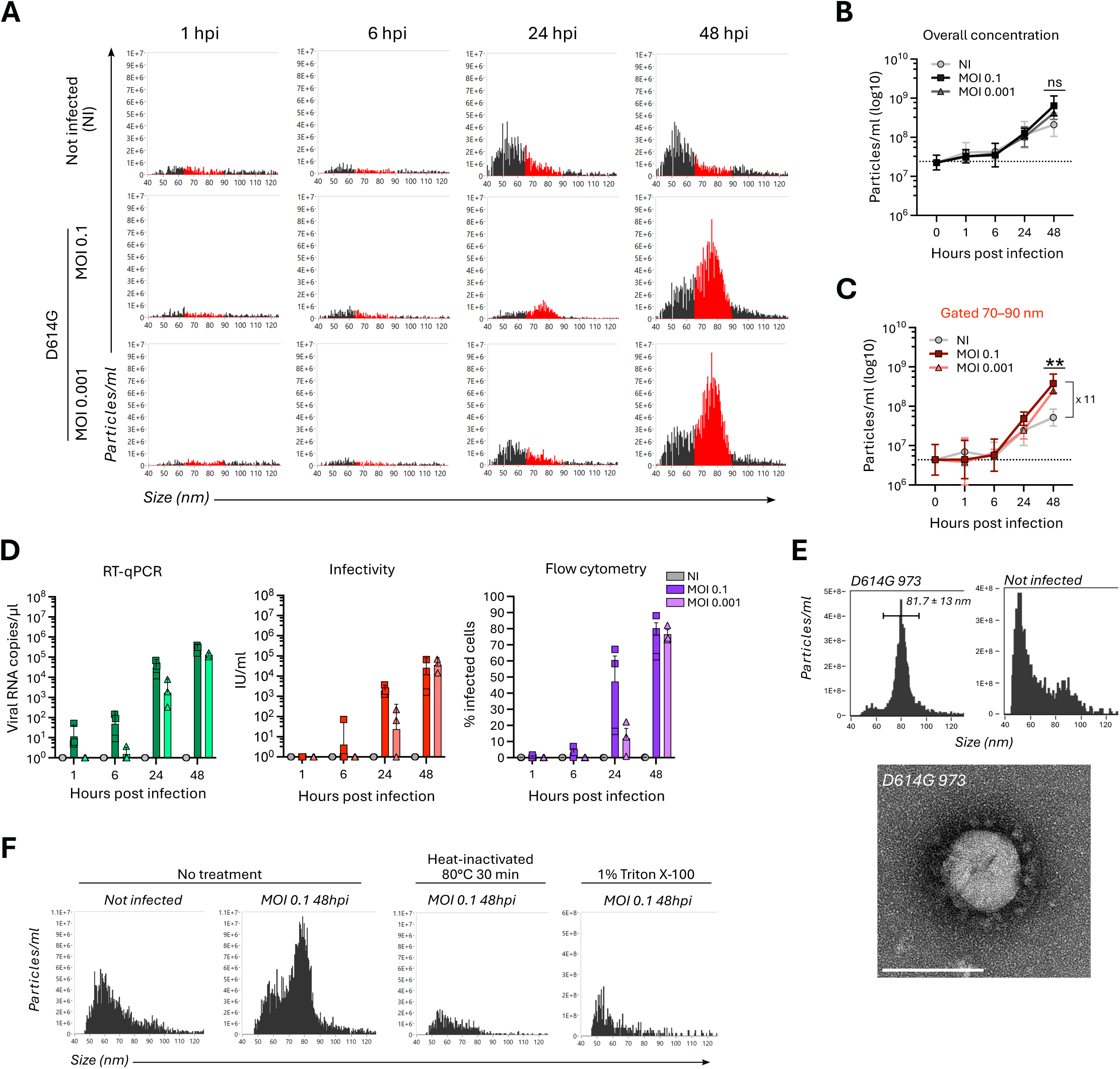
SARS-CoV-2 particles can be detected in infected cell supernatants without staining. (**A**) Size distribution plots of particles in supernatants from non-infected and SARS-CoV-2 D614G-infected VeroE6 cells (variant D614G at MOI 0.1 and 0.001 respectively) from different hours post infection (hpi). Red highlighted peaks are located between 70–90 nm. **(B and C)** Concentration measurements (particles/ml) of all events (B) and gated between 70–90 nm (C) from non-infected and D614G-infected VeroE6 supernatant. Dashed line indicates background measured at 0 hpi. Shown are the mean ± SD of 3 independent experiments. Statistical significance was determined by one-way ANOVA with Dunnett’s test for multiple comparisons. ns; not significant, **p < 0.01 **(D to F)** Quantification of viral RNA (RT-qPCR), infectious units per ml (IU/ml; S-Fuse) and percent infected cells (flow cytometry) from non-infected and D614G-infected VeroE6 supernatant. Shown are the mean ± SD of 3 independent experiments. **(G)** Size distribution plots of particles in the supernatant from non-infected and D614G-infected VeroE6 treated for heat-inactivation at 80°C for 30 min or treated with 1% Triton X-100. **(H)** Size distribution plots of particles and negative stain electron microscopy imaging from a SARS-CoV-2 viral stock (D614G) and supernatant (SN) from non-infected VeroE6 cells. Scale bars show 100 nm.

### Specific labelling of SARS-CoV-2 particles

Next, we established a procedure to fluorescently label SARS-CoV-2 particles **(Figure 2A)**. To stain the spike protein, RNA genome and lipid envelope, we tested the anti-spike antibody Tixagevimab conjugated to Dylight-488 (DL488), Syto24 and MemGlow-488, respectively **(Figure 2B)**. To reduce background fluorescence, we simply diluted the stained samples in PBS prior to acquisition. The unstained D614G sample revealed different background elevations depending on the dye **(Figure 2C)**. Tixagevimab-DL488 showed elevated background, likely due to unbound antibodies. Syto24 exhibited no background signal, probably because it requires double-stranded nucleic acid to activate its fluorescence. MemGlow-488 displayed no background signal but elevated noise, for which size and fluorescence are strongly associated. This is consistent with the nature of this dye, composed of fluorescent micelles that fuse with target membranes. Therefore, in the absence of targets, we directly detect the dye. In the supernatant of SARS-CoV-2-infected cells, the three dyes revealed distinct populations, always appearing at the expected position of viral particles **(Figure 2C)**. All events are stained with MemGlow-488, suggesting that all particles possess an envelope. In contrast, not all particles are stained with Syto24 or Tixagevimab. This might be due to decreased sensitivity of these stainings, or the lack of RNA or spike in some particles.

**Figure 2.**
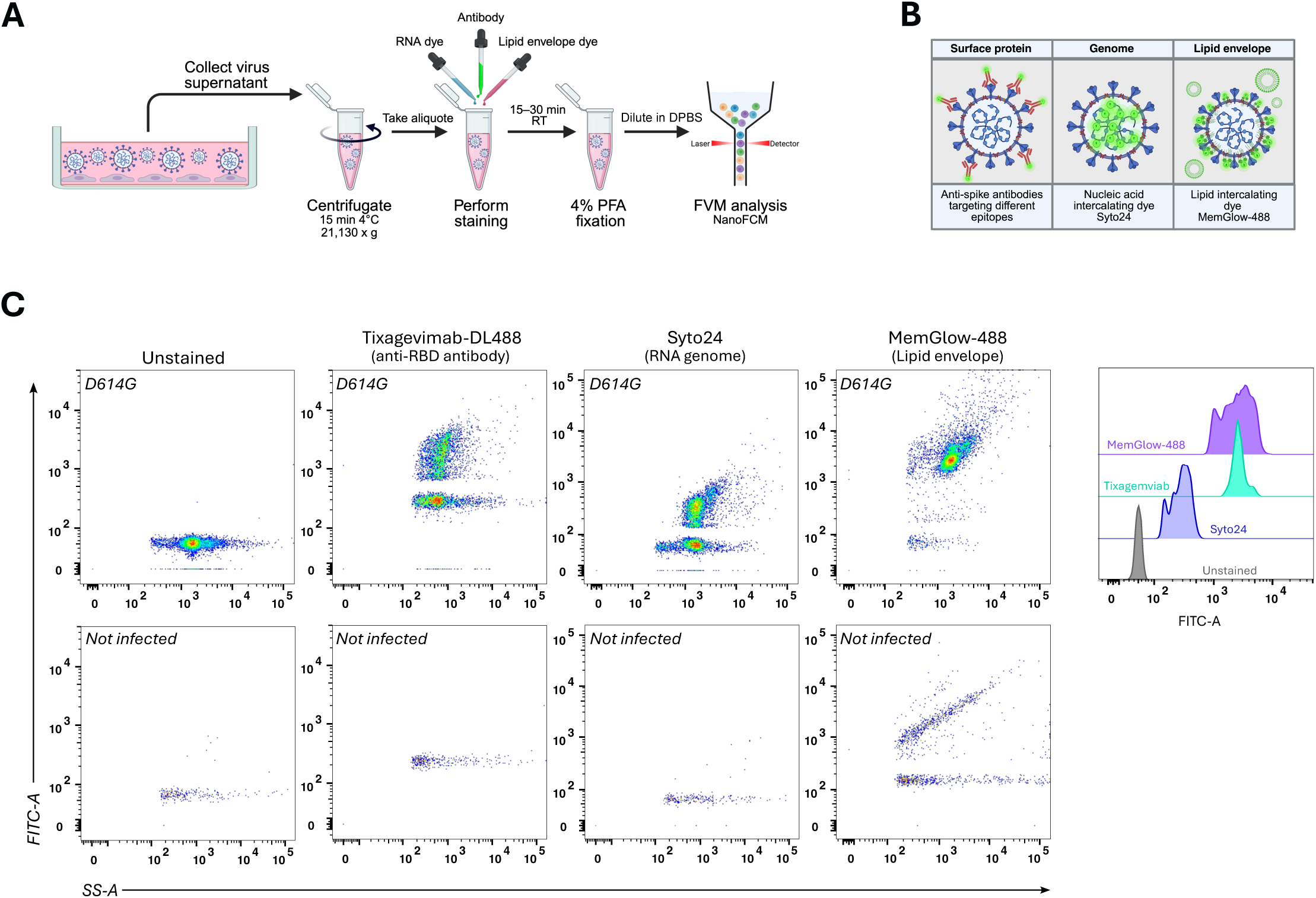
Procedure and labelling strategies for phenotyping SARS-CoV-2 particles. **(A)** Schematic representation of staining procedure for virus-containing supernatant. Supernatant from infected cells is harvested and centrifugated to remove cellular debris. Staining reagents are added to the sample, fixed with paraformaldehyde (PFA) and diluted in DPBS before being analyzed by flow virometry (FVM). **(B)** Graphical summary of different labelling techniques for targeting the SARS-CoV-2 spike protein via antibodies, the viral RNA genome via the nucleic acid intercalating dye Syto24 and the viral lipid envelope via the lipid intercalating dye MemGlow-488. **(C)** Dot plots and MFI of D614G stained with monoclonal antibody Tixagevimab (1 µg/mL; anti-RBD) conjugated to DyLight488 (DL488), with Syto24 (20 µM), with MemGlow-488 (200 nM), respectively.

Overall, these data demonstrate that fluorescent labeling of SARS-CoV-2 viral particles and subsequent analysis by FVM are feasible with both antibodies and dyes.

### Flow virometry enables single viral particles analysis

To determine whether SARS-CoV-2 particles aggregate, which could cause a swarming effect compromising single particle analysis and subsequent concentration and phenotypic characterization, we performed serial dilutions of D614G stained with Tixagevimab-DL488 and measured particle concentration and the MFI of spike-positive events between 70–90 nm **(Figure 3A and 3B)**. The particle concentration behaved linearly with the dilution, while the MFI remained stable, both of which indicate that single particles are interrogated **(Figure 3C)**. This limiting-dilution experiment also enabled estimation of the FVM limit of detection (LOD). We estimated it at approximately 10⁶ particles/mL when using both size and fluorescence for detection, compared to ∼10⁷ particles/mL for side scatter-based detection alone. Moreover, negative-stain transmission electron microscopy confirmed the presence of single SARS-CoV-2 particles without aggregation at approximately 100 nm in size **(Figure 3D)**.

**Figure 3.**
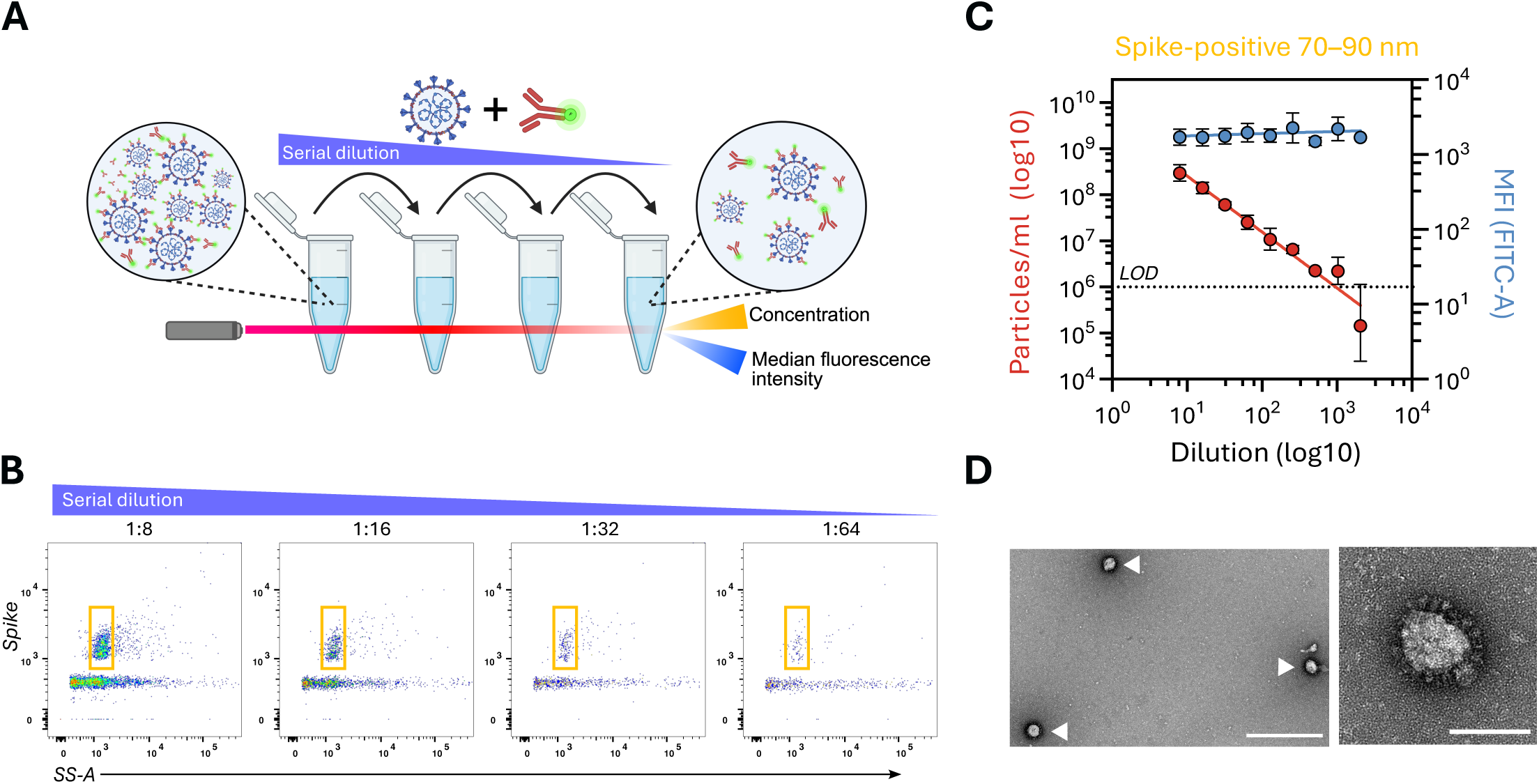
Flow virometry enables single SARS-CoV-2 viral particle analysis. **(A)** Schematic representation of serial dilution of virus-containing sample to infer swarming effect. **(B)** Representative dot plots of serially diluted D614G stained with Tixagevimab-DL488 (1 µg/mL). **(C)** Concentration and median fluorescence intensity (MFI) measurements of D614G spike-positive particles gated between 70–90 nm. Dashed line indicates the limit of detection (LOD). Red line indicates simple linear regression. Shown are the mean ± SD of 3 independent measurements. **(D)** Negative stain transmission electron miscopy images of D614G. White triangles highlight SARS-CoV-2 particles. Scale bars indicate 500 (left) and 100 (right) nm.

### The fluorescence intensity of spike is a better predictor of infectivity than particle concentration

Since crude viral preparations consist of a heterogeneous assemblage of defective and infectious viral particles, we wondered whether FVM could predict infectivity. For this, we took different D614G preparations, measured infectious titers by TCID50 and investigated particle concentration and spike MFI using Tixagevimab-DL488 **(Table 1)**. Preparations 1 and 2 showed the highest percentages of spike-positive events (77.8% and 66.9%, respectively), followed by preparations 7 (66.7%), 4 (54.0%), 3 (37.1%), 6 (25.0%) and lastly preparation 5 (16.7%) **(Figure 4A)**. The median concentration of spike-positive particles was more than 5,000-fold higher than that of infectious particles **(Figure 4B)**. Spearman correlation of particle concentration or spike MFI with infectious titer was not significant in either case, despite a trend toward improvement with spike MFI **(Figure 4C and 4D)**. When we compared infectious preparation 1 with the least infectious preparation 3 by transmission electron microscopy, we observed more particles with distinct spike protrusions on the surface, resembling SARS-CoV-2, whereas preparation 3 contained more extracellular vesicle-like objects **(Figure 4E and 4F)**. These data show that FVM enables assessment of viral particle quality together with concentration, providing information on the infectiousness of the viral population.

**Figure 4.**
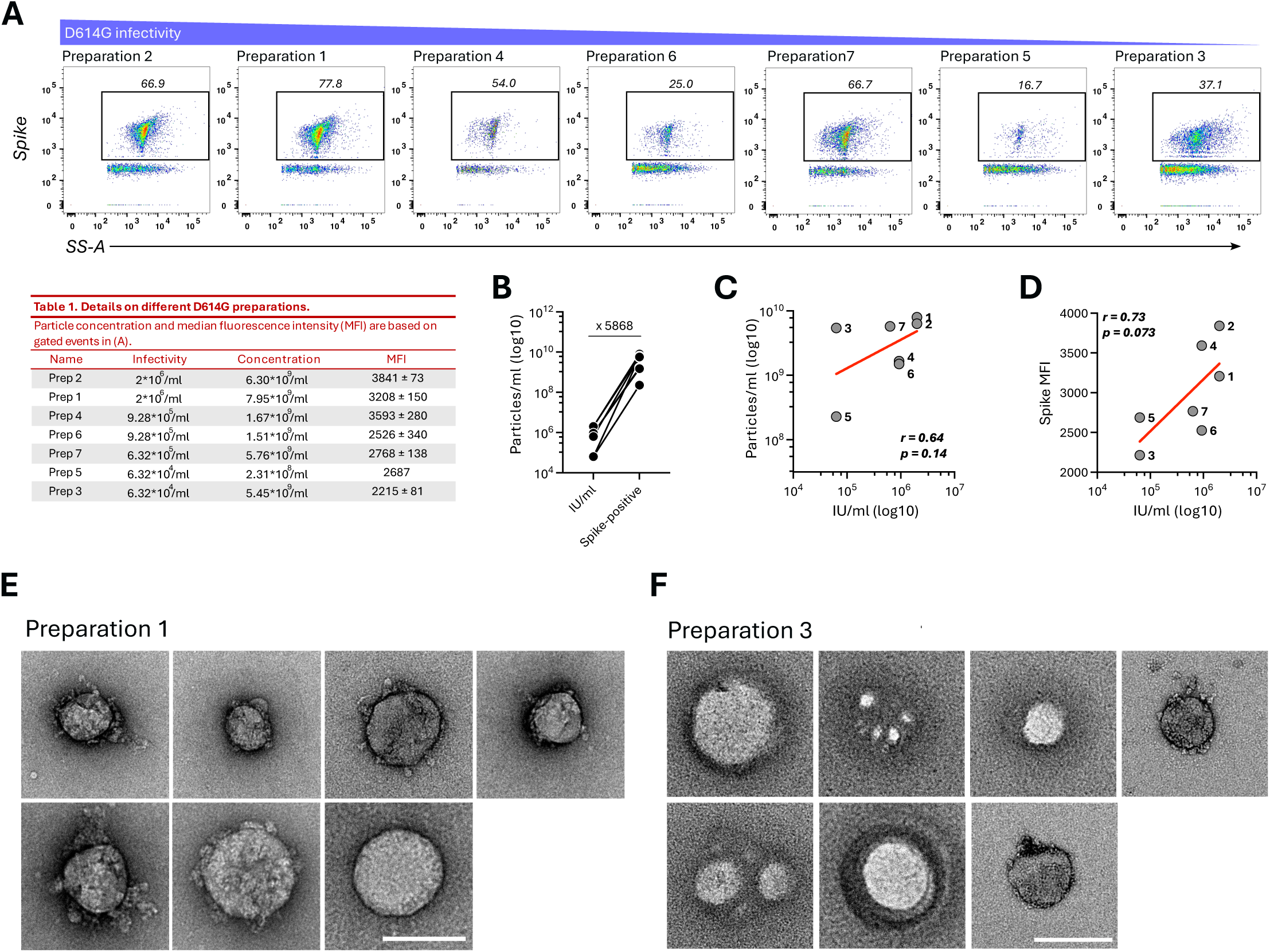
Spike MFI correlates better with infectivity than particle concentration. **(A)** Representative dot plots of seven different D614G preparations stained with Tixagevimab-DL488 (1 µg/mL). Table 1 illustrates various parameters from the different D614G preparations. **(B)** Concentration comparison between infectious (IU/ml) and spike-positive particles from all seven different D614G preparations. **(C and D)** Spearman correlation analyses between infectious particles (IU/ml) and all spike positive events or spike median fluorescence intensity (MFI). Gated spike-positive events are displayed in (A). Red line indicates simple linear regression analysis. **(E and F)** Negative stain transmission electron microscopy (TEM) of D614G preparation 1 and D614G preparation 3. Scale bar indicates 100 nm.

### Flow virometry enables the mapping of spike conformation at the single viral particle level

Next, we investigated whether FVM could detect changes in spike conformation using monoclonal antibodies as probes for distinct spike conformations. For this purpose, we utilized another instrument, the CytoFLEX nano as it allows multiplex staining. we confirmed that the double staining of viral RNA and spike with Syto24 and monoclonal antibody SA55 (anti-RBD), respectively, resembled the staining profile on the Flow NanoAnalyzer. Of note, the pipeline of the CytoFLEX nano requires additional gating strategies to minimize noise, such as gating on singlets and applying a cut-off threshold to the background **(Figure 5A)**. The staining profiles were similar between the two machines, but the CytoFLEX nano enabled the acquisition of up to 10 times more events. Most particles exhibited double stainings for viral RNA and spike. Compared to the Flow NanoAnalyzer, the CytoFLEX nano does not have an integrated tool for particle size estimation. Therefore, we calibrated the data using FCMpass software after acquisition. This resulted in a size estimation of 87.8 ± 5.61 nm, comparable to the Flow NanoAnalyzer and matching cryo-EM data for SARS-CoV-2 (82 ± 13 (69–95) nm; [5]. Next, we stained D614G with different anti-spike antibodies targeting the RBD (sotrovimab and SA55) and the fusion peptide (FP) within the S2 subunit (CoV44-62 and 76E1). While sotrovimab and SA55 exhibited distinct MFIs above the isotype control mGO53, binding by CoV44-62 and 76E1 was either weak or undetectable **(Figure 5B)**. The addition of ACE2 significantly reduced the binding MFI for both anti-RBD antibodies and increased it for both anti-FP antibodies compared to the condition without ACE2. This profile is consistent with receptor engagement on S and a transition from prefusion to post-fusion conformation in a subset of spike. Altogether, these data indicate that FVM enables detection of changes in the conformation of spike elicited by receptor engagement directly on single viral particles.

**Figure 5.**
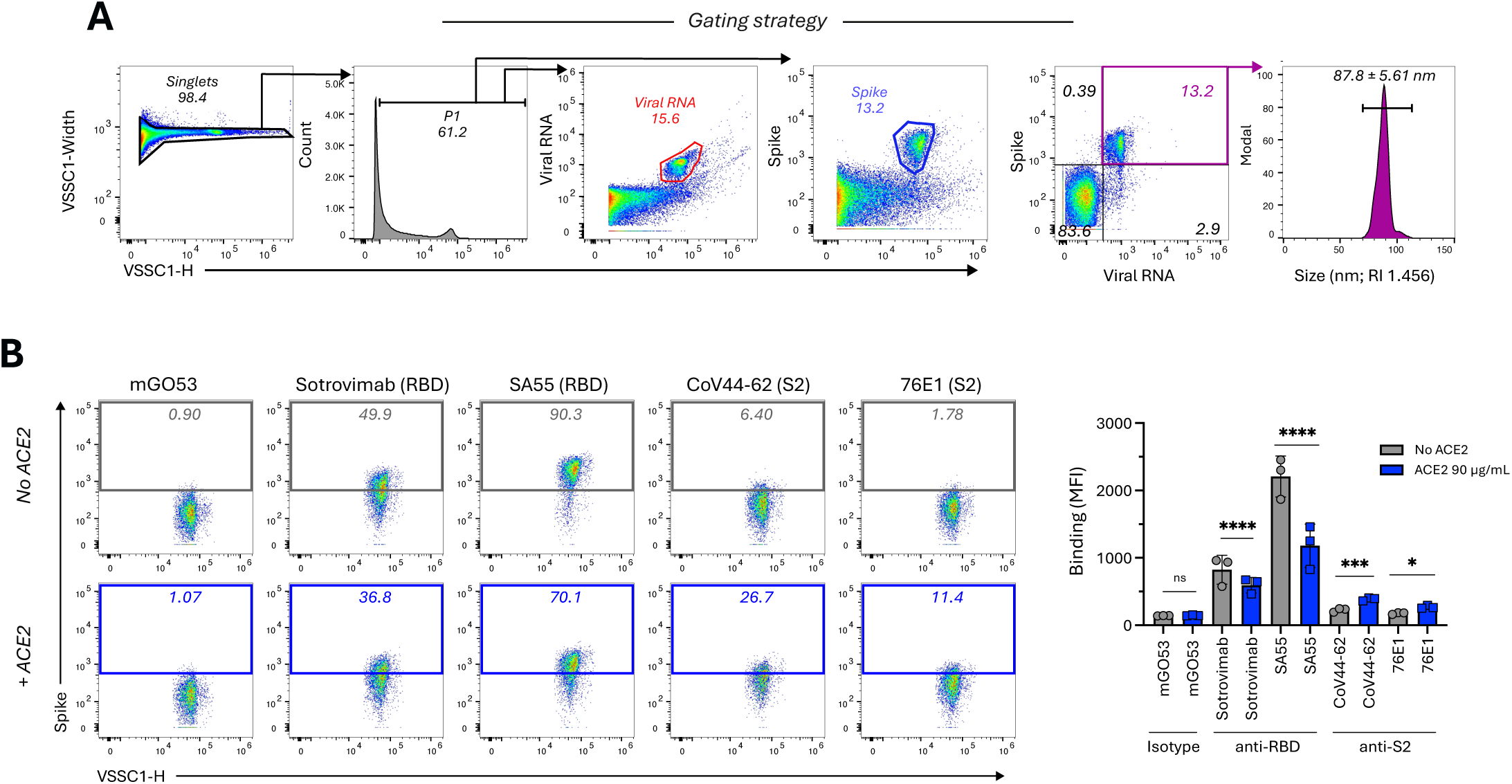
Probing of spike conformations on individual virion via antibodies. **(A)** Gating strategy on the Beckman CytoFLEX nano using D614G stained with Syto24 and SA55 (anti-RBD). To reduce background noise, first the singlets were selected, and the background was cut-off with the P1 gate, before the viral populations were gated. **(B)** Left panel: Representative dot plots of D614G 973 incubated with and without ACE2 (90 µg/mL) and stained with Syto24 and different mAbs (mGO53, sotrovimab, SA55, CoV44-62 and 76E1), respectively. Numbers indicate the percent of spike-positive events within the gate. Right panel: Antibody binding of respective mAbs. Shown are the mean ± SD of 3 independent measurements. Statistical significance was determined by two-way ANOVA with Sidak’s test for multiple comparisons. ns; not significant, *p < 0.05, **p < 0.01, ***p < 0.001, *****p < 0.0001

### Detection of SARS-CoV-2 particles in patient nasal swabs

We then asked whether we could detect SARS-CoV-2 particles in nasal swabs of PCR-confirmed COVID-19 patients using FVM. We used four nasal swabs collected in December 2023 for standard surveillance, consisting of one SARS-CoV-2-negative and three positive swabs (RT-qPCR; Ct 12, Ct 13 and Ct 16). The swabs were tested negative for other common respiratory viruses to exclude false-positive detection. After resuspension of the swabs in buffer, centrifugation to remove larger debris and dilution with DPBS, we stained each aliquot with SA55-DL488 due to its broad cross-reactivity with SARS-CoV-2 variants [20, 21] **(Figure 6A)**. The positive control virus XBB.1.5 exhibited a strong spike-positive population with a size of approximately 100 nm **(Figure 6B)**. While the negative swab showed no staining, the three SARS-CoV-2-positive swabs displayed spike-positive populations at approximately 100 nm. The swab with Ct 12 demonstrated the highest spike-positive events, followed by Ct 13 and lastly Ct 16. Altogether, these data demonstrate the capacity of FVM to detect SARS-CoV-2 viral particles directly in clinical samples, with minimal handling.

**Figure 6.**
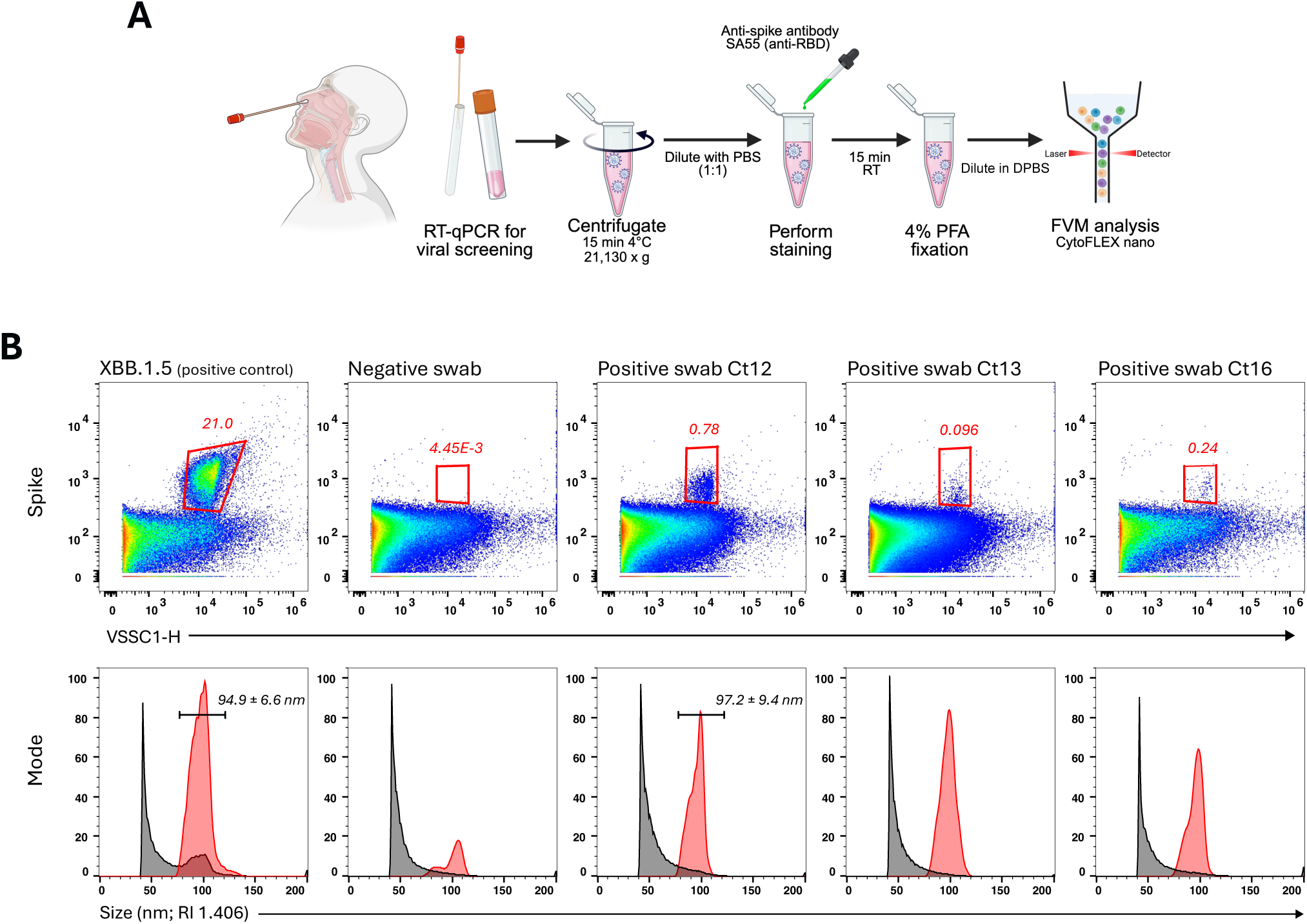
Detection of SARS-CoV-2 particles in nasal swabs from COVID-19 patients. **(A)** Schematic representation of nasal swab sample preparation and staining for SARS-CoV-2 particles. After nasal swab collection, the sample buffer was screened for different respiratory viruses including SARS-CoV-2 by RT-qPCR. SARS-CoV-2-positive samples, but negative for other respiratory viruses, were centrifugated before being diluted with PBS, stained with SA55 (anti-RBD), fixed in PFA and analyzed by FVM. **(B)** SA55 spike stainings of XBB.1.5 and SARS-CoV-2-negative and -positive nasal swabs with different amounts of viral RNA determined by RT-qPCR.

## Discussion

Flow virometry (FVM) has emerged as a powerful tool for interrogating viral heterogeneity at the single particle level, yet its application to respiratory viruses such as SARS-CoV-2 has remained underexplored. We established a robust framework for detecting and phenotyping individual SARS-CoV-2 particles directly from cell culture supernatants and patient nasal swabs, without laborious purification steps. By integrating label-free side scatter analysis with multiparametric fluorescence staining, we demonstrate that SARS-CoV-2 appears as discrete 70–100 nm particles that can be quantified and characterized for their lipid envelope, genomic RNA content, spike protein incorporation and conformation. These capabilities reveal previously inaccessible viral populations and provide quantitative metrics that correlate with infectious potential.

A key challenge in studying enveloped viruses is distinguishing *bona fide* virions from the abundant background of extracellular vesicles (EVs) and protein aggregates present in biological samples [22]. Our data indicate that while label-free FVM can detect SARS-CoV-2 progeny particles based on side scatter properties alone, this approach requires high viral titers to resolve particles above background. The delayed detection by FVM (48 hpi) compared to RT-qPCR or functional infectivity assays (6–24 hpi) reflects the sensitivity limitations of scatter-based detection in complex media, where cellular debris and EVs generate substantial signals due to physical and biochemical similarities. This limitation underscores that while label-free FVM offers convenient near-real-time monitoring of particle shedding, it is best suited for late-stage infection analysis or conditions where viral titers exceed 10^6^ particles/mL. The absence of a 70–90 nm peak in heat-inactivated or detergent-treated samples confirms that FVM specifically detects intact and enveloped viral particles.

To overcome sensitivity limitations and enable specific phenotyping, we developed complementary staining strategies targeting distinct viral components. We observed differential staining patterns, where all particles incorporated MemGlow-488 (envelope) while only subsets were positive for Syto24 (RNA) or anti-spike antibodies, providing insight into the heterogeneity of viral populations. The envelope staining confirms the presence of the lipid bilayer across the particle population, whereas the variable RNA and spike signals likely reflect the presence of defective viral particles lacking genomes or envelope glycoproteins, respectively. Such heterogeneity has profound implications for viral fitness and immune evasion, as non-infectious particles may act as decoys for neutralizing antibodies [23].

The improved signal-to-noise ratio achieved with anti-spike antibodies (particularly those targeting the RBD) compared to Syto24 is likely reflected by the surface accessibility of spike trimers. Conversely, the low signal intensity of Syto24 could be due to restricted regions that have higher-folded RNA structures coupled with lower nucleic acid content. This hypothesis is corroborated by previous work on human cytomegalovirus where Syto13 yielded stronger signals [24]. The linear relationship between particle concentration and fluorescence intensity upon serial dilution, confirmed by transmission electron microscopy, validates that FVM interrogates single, non-aggregated virions, a critical prerequisite for accurate quantification and functional correlation.

Quantitative analysis of different viral preparations revealed that median fluorescence intensity (MFI) of spike staining is a more robust predictor of infectious titer than particle concentration alone. This finding aligns with the requirement for spike-mediated receptor binding and membrane fusion during entry. However, the observed disconnection between total particle counts and infectivity (up to 5,000-fold more physical particles than infectious units) highlights the prevalence of defective or suboptimal particles in coronavirus populations. The correlation between spike MFI and infectivity suggests that FVM could serve as a rapid quality control tool for evaluating viral preparations or to enable the screening of antiviral strategies that target viral particles. Future studies should determine whether an optimal spike density exists for maximal infectivity, as suggested by work on other enveloped viruses [25, 26], and how spike incorporation varies across different SARS-CoV-2 variants or host cell types.

Our ability to monitor spike conformational changes using domain-specific monoclonal antibodies demonstrates that FVM can capture dynamic structural transitions on individual viral particles. The reciprocal binding patterns of RBD-targeting and fusion peptide-targeting antibodies upon ACE2 engagement (*i.e.* decreased RBD accessibility concomitant with increased fusion peptide exposure) provide direct evidence of prefusion-to-postfusion conformational switching at the single particle level. This capability is particularly relevant for understanding how mutations in emerging variants affect spike stability and fusogenicity, and for characterizing the mechanism of action of therapeutic antibodies or entry inhibitors.

The detection of SARS-CoV-2 particles in unpurified patient nasal swabs represents a significant step toward clinical translation. The inverse correlation between RT-qPCR Ct values and spike-positive event counts suggests that FVM could complement existing diagnostic strategies by providing a direct measure of intact viral particles rather than total nucleic acid. The ability to discriminate intact, RNA positive, spike-displaying particles from degraded viral material in clinical specimens could inform on transmissibility and antiviral therapy efficacy. However, the elevated autofluorescence and particulate background in clinical samples necessitate careful gating strategies and may limit sensitivity. Previous works have elaborated a pipeline for antibody-based detection of SARS-CoV-2 particles from patient saliva and nasal swabs [27, 28], further suggesting that direct quantification of viral particles in patient fluids might be a cost and time-effective way to diagnose respiratory viral infection.

Methodologically, our cross-platform validation using both the Flow NanoAnalyzer and CytoFLEX nano demonstrates the portability of our staining pipeline across instruments. The slight discrepancies in size estimation (70–100 nm across platforms versus 82 ± 13 nm by cryo-EM) highlight current limitations in flow virometry calibration, primarily coming from uncertainties regarding the refractive index (RI) of virions *versus* synthetic calibration standards. While recent software iterations utilizing an RI of 1.456 for enveloped viruses closely approximate cryo-EM measurements, precise determination of SARS-CoV-2 RI remains essential for standardizing size measurements across laboratories.

Several limitations warrant consideration. First, while our staining approach improves sensitivity, the absolute limit of detection of 10^6^ may preclude analysis of samples with low viral loads. Our patient cohort was small and retrospective. Therefore, prospective studies with larger sample sizes, longitudinal sampling, and correlation with viral culture are needed to validate FVM as a clinical diagnostic tool. Future applications of this technology could include characterizing the impact of humoral immunity on viral particle composition or studying the heterogeneity of shedding across various cell culture system and clinical respiratory samples.

In conclusion, we have established FVM as a versatile platform for the single particle analysis of SARS-CoV-2, enabling simultaneous quantification, phenotyping, and conformational interrogation of individual virions. This framework offers new opportunities to dissect the functional consequences of viral heterogeneity in coronavirus biology and holds promise for rapid, particle-based diagnostics and therapeutic monitoring.

## Acknowledgements

We thank members of the Virus and Immunity Unit for discussions and help in the preparation of this manuscript. We thank Jocelyne Creff and Fabienne Tzvetkov-Ricard for their technical and administrative assistance. We thank the entire PPU community of the Pasteur Paris-University for their continuous support. We thank Sylvie Van der Werf and the BioSPC doctoral school for the help during the thesis. We thank Arnaud Morris, Nadia Naffakh, Clarisse Berlioz-Torrent, Sarah Gallois-Montbrun and Julie Guignot for their participation to the thesis Committee of MJG. We thank Divya Unni and the Institut Pasteur grant office for their unvaluable assistance in funding acquisition. We thank N. Aulner and the UtechS Photonic BioImaging (UPBI) core facility (Institut Pasteur), a member of the France BioImaging network, for image acquisition and analysis. We thank the patients who participated to this study.

## Author contributions

Experimental strategy and design: M.J.G., T.B.

Laboratory experiments: M.J.G., P.H.C., A.C.C., A.D.C., A.F.R., C.P., F.G.B., I.S.

Cohort management and clinical research: D.V., H.P.

Key reagents and techniques: P.H.C., C.P., S.S., S.N., H.M.

Funding acquisition: S.S., S.N., H.M., O.S., T.B. Manuscript writing & editing: M.J.G., T.B.

## Data Availability

All data produced in the present study are available upon reasonable request to the authors

## Inclusion and diversity

We support inclusive, diverse, and equitable conduct of research.

## References

1. Sardanyés, J., et al., Quasispecies theory and emerging viruses: challenges and applications. npj Viruses, 2024. 2(1): p. 54.

2. Lippé, R., Flow Virometry: a Powerful Tool To Functionally Characterize Viruses. Journal of Virology, 2018. 92(3): p. e01765–17.

3. Fernandes, C., et al., Flow virometry: recent advancements, best practices, and future frontiers. J Virol, 2025: p. e0171724.

4. Zamora, J.L.R. and H.C. Aguilar, Flow virometry as a tool to study viruses. Methods, 2018. 134–135: p. 87-97.

5. Yao, H., et al., Molecular Architecture of the SARS-CoV-2 Virus. Cell, 2020. 183(3): p. 730–738.e13.

6. Harvey, W.T., et al., SARS-CoV-2 variants, spike mutations and immune escape. Nature Reviews Microbiology, 2021. 19(7): p. 409–424.

7. Ke, Z., et al., Structures and distributions of SARS-CoV-2 spike proteins on intact virions. Nature, 2020. 588(7838): p. 498–502.

8. Murigneux, E., et al., Proteomic analysis of SARS-CoV-2 particles unveils a key role of G3BP proteins in viral assembly. Nat Commun, 2024. 15(1): p. 640.

9. Buchrieser, J., et al., Syncytia formation by SARS-CoV-2-infected cells. Embo j, 2020. 39(23): p. e106267.

10. Planas, D., et al., Resistance of Omicron subvariants BA.2.75.2, BA.4.C and BQ.1.1 to neutralizing antibodies. bioRxiv, 2022.

11. Planas, D., et al., Considerable escape of SARS-CoV-2 Omicron to antibody neutralization. Nature, 2022. 602(7898): p. 671–675.

12. Bruel, T., et al., Antiviral activities of sotrovimab against BQ.1.1 and XBB.1.5 in sera of treated patients. medRxiv, 2023.

13. Planas, D., et al., Distinct evolution of SARS-CoV-2 Omicron XBB and BA.2.86/JN.1 lineages combining increased fitness and antibody evasion. Nat Commun, 2024. 15(1): p. 2254.

14. Monel, B., et al., Release of infectious virus and cytokines in nasopharyngeal swabs from individuals infected with non-alpha or alpha SARS-CoV-2 variants: an observational retrospective study. EBioMedicine, 2021. 73: p. 103637.

15. Welsh, J.A. and J.C. Jones, Small Particle Fluorescence and Light Scatter Calibration Using FCMPASS Software. Current Protocols in Cytometry, 2020. 94(1): p. e79.

16. Welsh, J.A., J.C. Jones, and V.A. Tang, Fluorescence and Light Scatter Calibration Allow Comparisons of Small Particle Data in Standard Units across Different Flow Cytometry Platforms and Detector Settings. Cytometry Part A, 2020. 97(6): p. 592–601.

17. Welsh, J.A., et al., FCMPASS Software Aids Extracellular Vesicle Light Scatter Standardization. Cytometry Part A, 2020. 97(6): p. 569–581.

18. Welsh, J.A., et al., A compendium of single extracellular vesicle flow cytometry. J Extracell Vesicles, 2023. 12(2): p. e12299.

19. Planchais, C., et al., Potent human broadly SARS-CoV-2-neutralizing IgA and IgG antibodies effective against Omicron BA.1 and BA.2. J Exp Med, 2022. 219(7).

20. Planas, D., et al., Escape of SARS-CoV-2 variants KP1.1, LB.1 and KP3.3 from approved monoclonal antibodies. bioRxiv, 2024: p. 2024.08.20.608835.

21. Cao, Y., et al., Rational identification of potent and broad sarbecovirus-neutralizing antibody cocktails from SARS convalescents. Cell Reports, 2022. 41(12): p. 111845.

22. Zhou, Y., R.P. McNamara, and D.P. Dittmer, Purification Methods and the Presence of RNA in Virus Particles and Extracellular Vesicles. Viruses, 2020. 12(9).

23. Rezelj, V.V., L.I. Levi, and M. Vignuzzi, The defective component of viral populations. Curr Opin Virol, 2018. 33: p. 74–80.

24. Bokun, V., et al., Nano-Flow Cytometry-Guided Discrimination and Separation of Human Cytomegalovirus Virions and Extracellular Vesicles. J Extracell Vesicles, 2025. 14(5): p. e70060.

25. Monticelli, S.R., et al., An increase in glycoprotein concentration on extracellular virions dramatically alters vaccinia virus infectivity and pathogenesis without impacting immunogenicity. PLoS Pathog, 2021. 17(12): p. e1010177.

26. Gaudin, R. and N.S. Barteneva, Sorting of small infectious virus particles by ffow virometry reveals distinct infectivity profiles. Nature Communications, 2015. 6(1): p. 6022.

27. Hussain, R., et al., Small form factor flow virometer for SARS-CoV-2. Biomed Opt Express, 2022. 13(3): p. 1609–1619.

28. Alharbi, N.K., et al., Development and Evaluation of Enzyme-Linked Viral Immune Capture Assay for Detection of SARS-CoV-2. Frontiers in Bioengineering and Biotechnology, 2022. Volume 10 - 2022.

29. Pinto, D., et al., Cross-neutralization of SARS-CoV-2 by a human monoclonal SARS-CoV antibody. Nature, 2020. 583(7815): p. 290–295.

30. Zost, S.J., et al., Potently neutralizing and protective human antibodies against SARS-CoV-2. Nature, 2020. 584(7821): p. 443–449.

31. Dacon, C., et al., Broadly neutralizing antibodies target the coronavirus fusion peptide. Science, 2022. 377(6607): p. 728–735.

32. Sun, X., et al., Neutralization mechanism of a human antibody with pan-coronavirus reactivity including SARS-CoV-2. Nature Microbiology, 2022. 7(7): p. 1063–1074.

